# PLWHA’s Perspective on Community Stigma for Getting Social Support and Improving Life Quality in the Digital Era

**DOI:** 10.1101/2022.01.17.22269334

**Authors:** Rio Ady Erwansyah, Moses Glorino Rumambo Pandin

**Affiliations:** Student of Doctoral Program in Nursing, Faculty of Nursing, Airlangga University; Faculty of Humanities, Airlangga University

**Keywords:** Social Support, Stigma, PLWHA/ODHA, HIV, AIDS

## Abstract

Community stigma against HIV/AIDS is still a problem that must be faced by people living with HIV/AIDS (PLWHA). The existence of stigmatization and discrimination received by PLWHA causes the lack of social support they get. Social support is very important for PLWHA to improve their quality of life. In this digital era where the development of technology and social media is an alternative for PLWHA to get social support online through their community, by sharing information and experiences. This systematic review was conducted through Scopus, Science Direct, ProQuest, and SAGE. The selection of articles followed inclusion criteria, including articles published in the last five years and published in English and discussing perspectives on stigma. Articles published not in English and articles with unclear literature reviews were excluded from this study. The PRISMA flow chart and JBI assessment checklist were used to assess the risk of bias and article quality. 20 relevant articles will be reviewed. The results of the study found that in this digital era PLWHA can take advantage of technological developments and social media to improve their quality of life by getting social support through the online communities they follow.

## 1. Background

Stigma against HIV/AIDS remains a significant barrier to public health worldwide and a major barrier to HIV/AIDS treatment, prevention, and support. Stigma against people living with HIV/AIDS (PLWHA) leads to reduced economic income and increased isolation from family and friends regarding their HIV status^1^. PLWHA often experience physical, psychological, social, and spiritual suffering, which harms their quality of life ^2-3^

PLWHA face several difficulties when they try to achieve a satisfactory quality of life, from disturbances in their life history, disturbances in interpersonal and work relationships that can cause social isolation, and worsen social relationships that can harm their mental and physical health^4^. In this digital era, it is still difficult for them to adapt to their current status changes regarding the diagnosis they receive. Stigma and community discrimination against HIV/AIDS are still considered a major problem in the survival of PLWHA, making it difficult for them to get social support from both the community and family. Whereas social support from the community, especially families, can help PLWHA improve their quality of life and assist in the acceptance and disclosure of their serostatus.

## 2. Objective

The purpose of this systematic review is to explore the perspective of PLWHA on community stigma in obtaining social support and improving their quality of life in the digital age.

## 3. Methodology

This research is a review of articles conducted to find out the perspective of PLWHA in the digital era. The checklist and flow chart of reporting items for systematic review (PRISMA) was used to present the results of the systematic review. We entered electronic databases to find relevant articles such as Scopus, Science Direct, ProQuest, and SAGE. The literature search process was determined in a recent study of the last five years. There are no regulatory restrictions, but only articles published in English are reviewed. The feasibility of this study was assessed using the PICOT framework as below.

### Bias Risk

The risk of bias can be reduced by using critical assessment tools. The review used a critical assessment tool from the Joanna Briggs Institute (JBI). The JBI assessment tool is a checklist questionnaire with a list of required items, things to do, and points to consider. Each research design has different questions. Researchers must assess the articles that have been selected. The scoring results come from low risk, medium risk, high risk, or unclear risk.

Inclusion and exclusion criteria: the articles used, filtered, and selected based on the inclusion criteria that have been set with the provisions of 5 years from 2017 to 2021, contain a discussion of perspectives on stigma. Articles that pass the inclusion criteria are then eliminated by the exclusion criteria. Exclusion criteria were referred to when the article was not written in English, the article was more than five years old, and the literature had an unclear review. The data that has been obtained is reviewed and then selected to be further grouped and discussed based on the points

## 4. Result

An initial keyword search of the literature yielded 395 articles (59 from Scopus, 87 from Science Direct, 218 from ProQuest, and 31 from SAGE). After a review, 125 articles were eliminated because the population did not match the research objectives, the articles did not use English, and were not open access. Then 169 articles were excluded based on the last five years and did not include a literature review, and 101 complete articles that were assessed for eligibility were eliminated 81 articles because the articles did not discuss perspectives and the literature had an unclear review so that 20 articles were obtained that were following the feasibility of the study.

**Figure 1.**
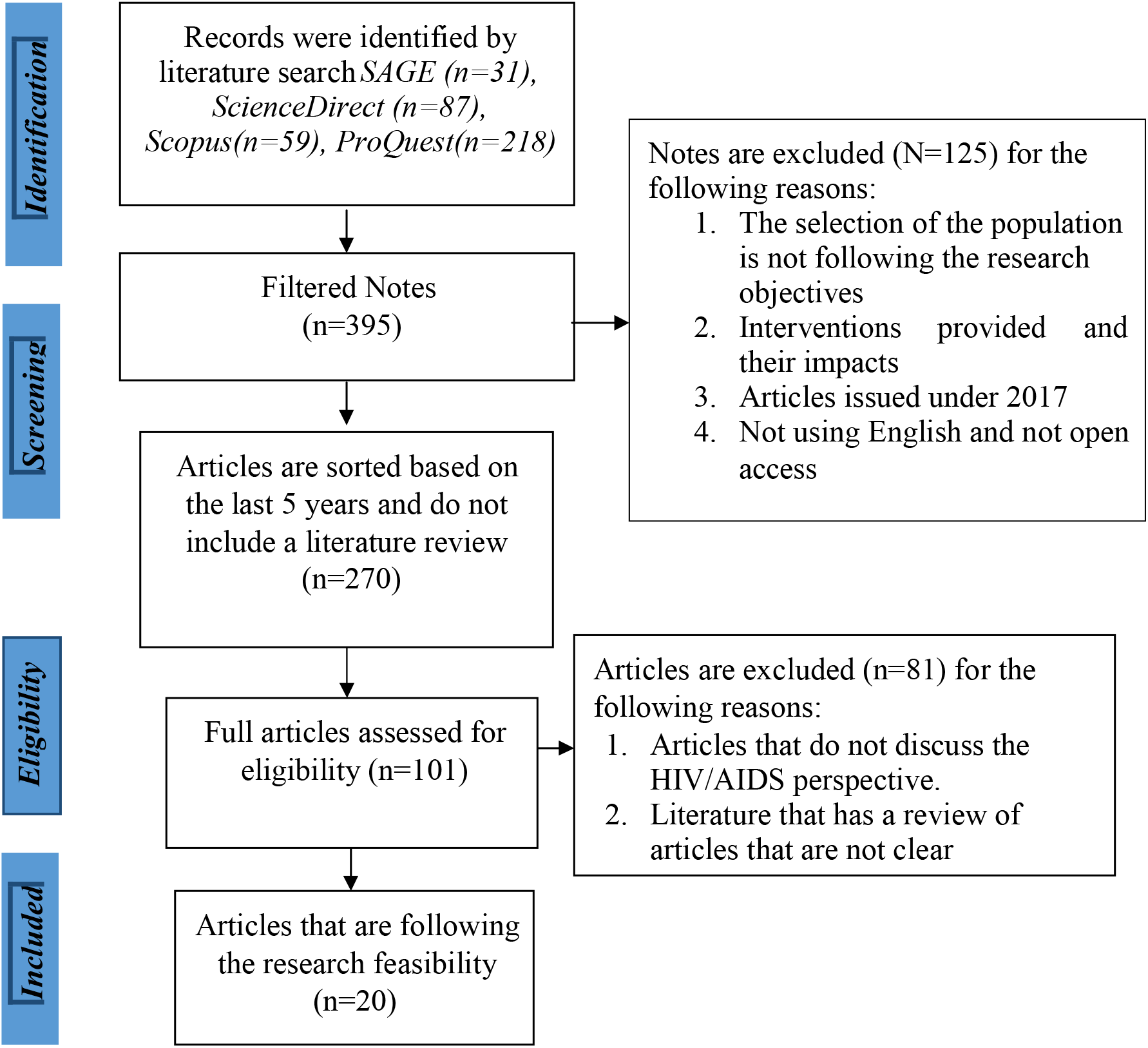
The results of the literature selection are summarized in the PRISMA flow diagram.

The risk of bias in the articles that we found can be seen in the following table:

The Joanna Briggs Institute (JBI) Critical Assessment was used to assess and analyze the methodological quality of the articles obtained (n = 20). As agreed by the researcher himself that the final score of the methodological quality assessment if it reaches a minimum of 75% meets the critical assessment criteria, the article will be included for further data synthesis. All articles (n = 20) at the last screening achieved a score higher than 75% so they were ready for data synthesis.

Based on table 2, 14 articles were assessed for risk of bias using the JBI Critical Assessment Checklist for Qualitative Studies and the results were: a score of 90% (n=10 articles) (N. O’Brien et al., 2017, Zepeda KGM et al.., 2019, Ahmed et al., 2019, Ernawati et al., 2021, Fauk et al., 2021, K. Srikanth Reddy, Seema Sahay, 2018, Jesus GJ, et al., 2017, Rutakumwa et al., 2021, Cherry S. at al., 2018, NS Perry et al., 2017), score 80% (n=4 articles) (Haruna T. et al., 2021, M. Abboah-Offei at al., 2019, Coelho B, Meirelles BHS., 2019, Rice et al., 2018)

4 articles were assessed for risk of bias using the JBI critical assessment checklist for the Cross-Sectional Study and the results were: a score of 87.5% (n=2 articles) (X. Han et al., 2018, Xie, Fie, et al., 2019), score 75% (n=2 articles) (CR Fumaz et al., 2019, Mehraeen et al., 2021).

2 articles were assessed for risk of bias using the JBI critical appraisal checklist for a cohort study and the results were: a score of 81.8% (n=2 articles) (Giraud, J.S, et al., 2020, S.M. Rein et al., 2021). Based on the identification in table 1, 20 articles were assessed for risk of bias using the JBI critical assessment checklist for the design of Cross-Sectional Study, Cohort Study, Prevalence Study, and QuasiExperimental Study. The results found were 20 articles had scores above 75%.

**Table 1.** Picot Framework.

| **PICOT Framework** | **Inclusion Criteria** |
| --- | --- |
| Population | PLWHA |
| Intervention | Explore experiences and perspectives |
| Comparator | No comparator |
| Output | PLWHA perspective |
| Time | 2017-2020 |

Articles are identified with the keywords “perspective, PLWHA, HIV, AIDS”.

**Table 2.**
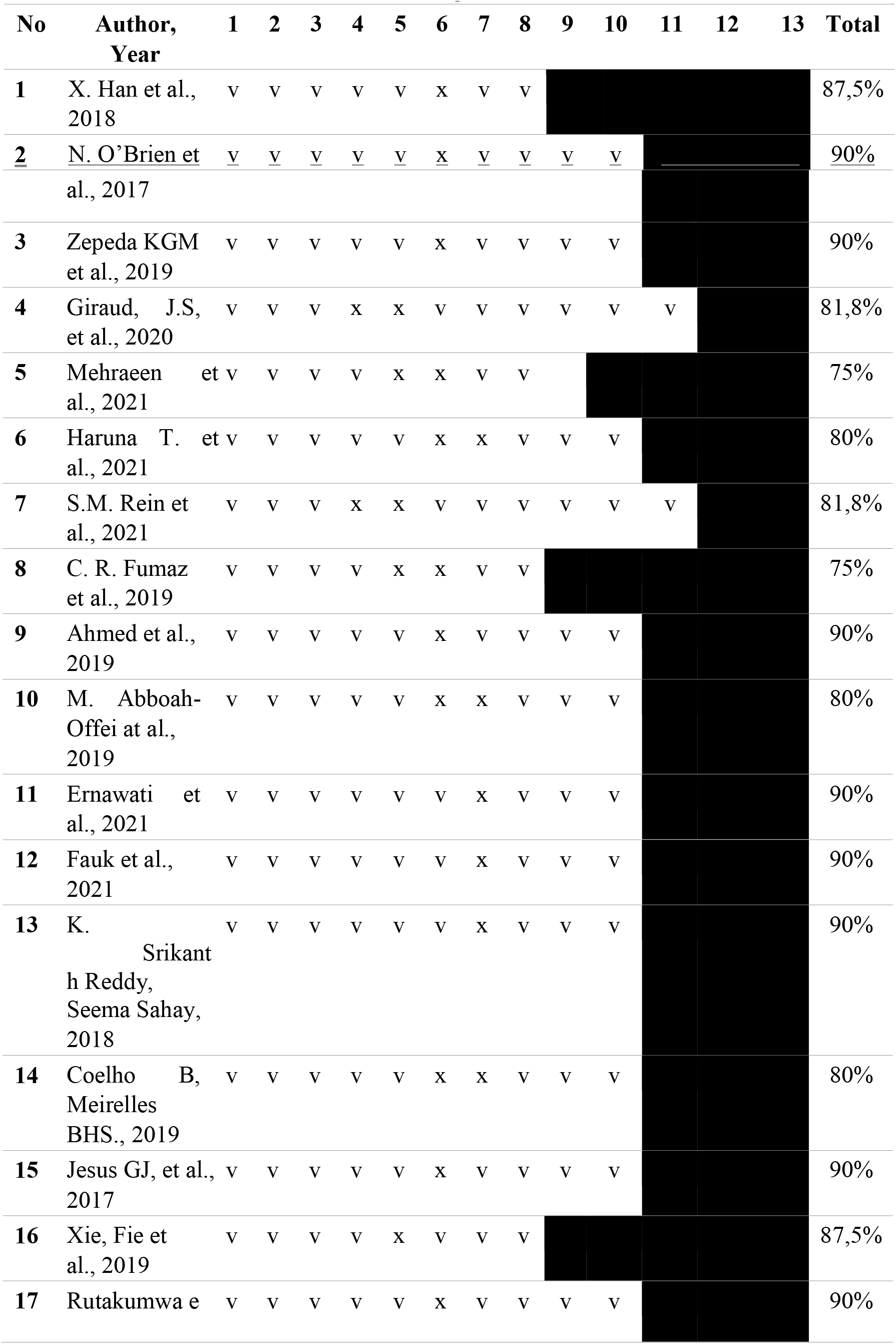
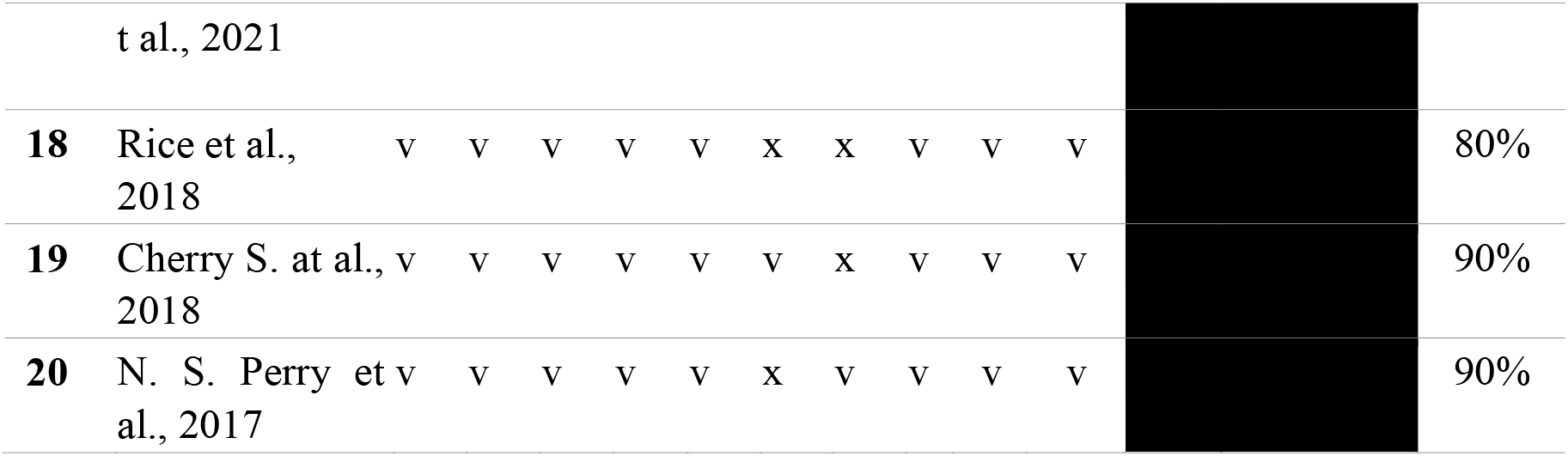
Risk assessment bias using the JBI critical assessment checklist

This systematic review was heterogeneous, with 14 articles using qualitative methods in the study, 4 cross-sectional articles, and 2 cohort articles. Results from selected articles using single and combined survey interventions. The sample varies because the journals selected are heterogeneous. The types of interventions provided are described in the following table:

1. Weibo friends with benefits for people living with HIV/AIDS? The implications of Weibo use for enacted social support, perceived social support, and health outcomes With the development of social media. Online support is becoming increasingly important for people with health problems such as HIV/AIDS to seek social support. Where the platform generates discussions on various health topics and becomes a place to share information and experiences ^5-6^
2. Envisioning Women-Centered HIV Care: Perspectives from Women Living with HIV in Canada A women-centered approach to HIV care is critical to guiding policy and practice to improve care and outcomes for women living with HIV ^7^.
3. Management of nursing care in HIV/AIDS from a palliative and hospital perspective In the context of palliative care, actions to improve comfort and quality of life are emphasized in the physical, socio-cultural, and psycho-spiritual fields in an altruistic way, to put oneself in the position of others to jointly try to understand the suffering of patients and families ^8^
4. De-simplifying single-tablet antiretroviral treatments for cost savings in France: From the patient perspectives to a 6-month follow-up on generics Simplification of ART treatment to generic drugs allows cost savings in health care costs ^9^
5. Investigating the contributing factors to HIV/AIDS infection from the perspective of HIV-infected patients An important cause of the spread of HIV is the government’s weakness in public health education, family and individual factors ^10^
6. Factors hindering the integration of care for non-communicable diseases within HIV care services in Dar es Salaam, Tanzania: The perspectives of health workers and people living with HIV NCD service bundling in HIV care and treatment clinics has adequate treatment adherence support to improve quality of life, close monitoring of biomarkers that can predict the development of NCDs and social support providers ^11^
7. Prospective association of social circumstance, socioeconomic, lifestyle, and mental health factors with subsequent hospitalization over 6-7 year follow up in people living with HIV Current markers of socioeconomic disadvantage (non-employment, insecure housing situations, and financial difficulties) are strong predictors of hospitalization among PLWHA ^12^
8. Health-related quality of life of people living with HIV infection in Spain: a gender perspective The stereotypic differences associated with each sex may explain the perceived differences in quality of life. Men adopt healthy behaviors, helping to improve physical and mental health, emotional status, and a better quality of life through physical exercise. While women appear to be affected by their quality of life by personal experiences of rejection, receiving social stigma from various community sources affects their physical health and coping strategies used in response to HIV-related stress ^13-14^
9. Facilitators and Barriers Affecting Adherence Among People Living With HIV/AIDS: A Qualitative Perspective Quality nursing outcomes depend on patient medication adherence. Lack of social support and poor doctor-patient communication often force patients to bear the burden of illness that affects their social and psychological well-being ^15^
10. How can we achieve person-centered care for people living with HIV/AIDS? A qualitative interview study with healthcare professionals and patients in Ghana PCC as care delivered in the community, free from stigma and discrimination, that addresses financial, human, and time resource issues ^16^
11. Life Experiences in Parenting: The Perspective of Women with HIV-AIDS The form of hope for negative HIV status from loved ones, the lamentation of wives with PLWHA status in the hope that they will remain healthy and hope that their children will not experience the same disease^17^
12. HIV Stigma and Discrimination: Perspectives and Personal Experiences of Healthcare Providers in Yogyakarta and Belu, Indonesia Stigma and discrimination are major challenges facing PLWHA globally because of their HIV status. Lack of knowledge about HIV, fear of being infected, personal values, religious thoughts, and socio-cultural values and norms are the driving forces for HIV-related stigma^18^
13. Advocacy for HIV/TB co-infection collaborative policy and service delivery in India: a civil society perspective HIV prevention is a priority for TB control, and TB care and prevention is a priority for HIV/AIDS programs ^19^
14. Care sharing for people with HIV/AIDS: a look targeted at young adults The Health Care Network (HCN) is an organization that regulates health services, including technology, which are interconnected by support systems, seeking to ensure comprehensive care ^20^
15. Difficulties of living with HIV/Aids: Obstacles to quality of life The impact of living with a chronic disease that still carries a lot of stigma and prejudice is the biggest obstacle between these individuals and their quality of life^21^
16. Social Capital Associated with Quality of Life among People Living with HIV/AIDS in Nanchang, China Bonding social capital score has a positive effect on mental health summary and physical health summary^22^
17. Stakeholders’ perspectives on integrating the management of depression into routine HIV care in Uganda: qualitative findings from a feasibility study Integrating depression management into routine HIV care is acceptable among key stakeholders^23^
18. Perceptions of Intersectional Stigma among Diverse Women Living with HIV in the United States Women living with HIV experience interrelated forms of stigma. The complex social environment in which HIV-positive women navigate shapes their realities of life, life opportunities, and well-being ^24^
19. “My determination is to live”: Narratives of African-American Women Who Have Lived with HIV for 10 or More Years Public health programs for long-term survivors, including ways to promote resilience, create opportunities for meaningful work, build skills and disclosure ^25^
20. Learning to address multiple syndemics for people living with HIV through client perspectives on CBT Future psychotherapeutic interventions for HIV syndromes and treatments address complex issues comprehensively ^26^

| No | Article Title | Intervention | Result |
| --- | --- | --- | --- |
| 1 | Weibo friends with benefits for people living with HIV/AIDS? The implications of Weibo use for enacted social support, perceived social support, and health outcomes | Filling out online questionnaires for HIV/AIDS positive Weibo users | Better socioeconomic conditions and poorer physical conditions were associated with higher PSS-ON rates. PLWHA experiences a higher level of social support via Weibo than offline. |
| 2 | Envisioning WomenCentered HIV Care: Perspectives from Women Living with HIV in Canada | Conduct focus group discussions with semistructured interviews to listen to the experiences of women living with HIV before shifting to women's perspectives on a women-centered | Women envision the 3 main characteristics of women-centered HIV care: coordinated and integrated services that address HIV and women's health care priorities that protect against exclusion from |
|  |  | approach to HIV care | treatment because of stigma, care that recognizes and responds to structural barriers, and cares that encourages peer support and leadership. same age |
| 3 | Management of nursing care in HIV/AIDS from a palliative and hospital perspective | Conduct semi-structured interviews in two sample groups with 17 participants | Generating an understanding of the relationship between nurse interactions in the management of nursing care for people who have clinical complications caused by AIDS and its relationship to palliative care |
| 4 | De-simplifying singletablet antiretroviral treatments for cost savings in France: From the patient perspectives to a 6-month follow-up on generics | 98 patients received a two-tablet treatment regimen containing the generic drug. 60 received Eviplera® and 38 received Triumeq®, six months later, a second survey was administered to patients | 45 patients received treatment that did not refuse simplified, 37 refused and 16 did not decide and followed the decision offered by their doctor |
| 5 | Investigating the contributing factors to HIV/AIDS infection from the perspective of HIV-infected patients | 150 patients with HIV were interviewed and filled out a medication adherence questionnaire and a mental health standard questionnaire | The results revealed that the main cause of HIV infection in men was an injection of drugs and in women was sexual intercourse with an infected individual. |
| 6 | Factors hindering the integration of care for non-communicable diseases within HIV care services in Dar es Salaam, Tanzania: The perspectives of health workers and people living with HIV | A guided checklist was observed for 5 days at the HIV care and treatment clinic on 130 PLWHA who provided PTM treatment services. Then conducted interviews with 5 CTC managers, 9 health care providers, and 27 PLWHA | There is a positive attitude between PLWHA and health workers towards the integration of noncommunicable disease services in HIV care and treatment clinics. This is due to the perceived benefits associated with the integration of PTM services in one clinic |
| 7 | Prospective association of social circumstance, socioeconomic, lifestyle, and mental health factors with subsequent hospitalization over 6-7 | Conducted the ASTRA study by recruiting PLWHA from 8 HIV outpatient clinics with a total of 798 participants and asking them to fill | Socio-economic losses, social conditions, evidence of mental health disorders, and adverse lifestyle factors are hospitalization for PLWHA |
|  | year follow up in people living with HIV | out a baseline questionnaire from February 2011 to June 2018 |  |
| 8 | Health-related quality of life of people living with HIV infection in Spain: a gender perspective | Evaluated gender differences in healthrelated quality of life in 744 PLHIV drawn from 32 Spanish hospitals offering HIV care, and participants were asked to complete the study assessment protocol with the supplied questionnaire. | Women who showed more emotional distress and negative moods related to their HIV condition used less positive coping strategies and experienced more rejection and stigma than men. |
| 9 | Facilitators and Barriers Affecting Adherence Among People Living With HIV/AIDS: A Qualitative Perspective | Conduct in-depth interviews with 18-yearold patients diagnosed with HIV/AIDS about medication adherence | Anxiety and stigma, the negative perceived image of HIV diagnosis, poor support from the community, and side effects of treatment are factors for treatment nonadherence. |
| 10 | How can we achieve person-centered care for people living with HIV/AIDS? A qualitative interview study with healthcare professionals and patients in Ghana | Conducted semi-structured interviews with PLWHA and health professionals at the community clinic aimed at participants' perspectives on their illness, access to care, involvement in decisionmaking, their symptoms, concerns, and preferences for care. | This study provides new insights into what patientcentered care (PCC) is all about. People with HIV understand PCC as care that involves them in their care decisions. |
| 11 | Life Experiences in Parenting: The Perspective of Women with HIV-AIDS | Conducted interviews with wives who were diagnosed with HIV/AIDS and registered by the HIV-AIDS program at the Serang City Health Center, to explore the phenomenon of mothers with HIV/AIDS in caring for their children. | The results showed that the mother wanted her child or husband to have a negative HIV status. |
| 12 | HIV Stigma and Discrimination: Perspectives and Personal Experiences of | Conducted interviews with 92 PLWHA and 20 health care providers regarding stigma and | Perspectives and experiences of health care providers regarding HIV stigma and discrimination |
|  | Healthcare Providers in Yogyakarta and Belu, Indonesia | discrimination against PLWHA | still occur in families and communities. Reflected in negative labeling by others |
| 13 | Advocacy for HIV/TB co-infection collaborative policy and service delivery in India: a civil society perspective | Conducted interviews with 26 participants consisting of representatives of advocacy organizations, health activists and PLWHA | Civil society prioritizes HIV/TB coinfection is an important public health issue and advocates identifying common indicators of coinfection for better program management |
| 14 | Care sharing for people with HIV/AIDS: a look targeted at young adults | Conducting interviews with 19 participants with HIV/AIDS who were divided into 2 groups was shown to understand the sharing of care for PLWHA | City governments have transitioned into a model of health care for people with HIV/AIDS, whose care is shared within the health care network. |
| 15 | Difficulties of living with HIV/Aids: Obstacles to quality of life | Conducted interviews with 26 PLWHA about quality of life | There are difficulties faced by PLWHA. The main difficulties cited are prejudice experienced in family and social contexts, managing affective and sexual partnerships, and achieving quality of life |
| 16 | Social Capital Associated with Quality of Life among People Living with HIV/AIDS in Nanchang, China | 225 PLWHA in Nanchang, China was asked to fill out a questionnaire to identify social capital factors that identify the quality of life in PLWHA | People living with HIV have low social capital and poor quality of life in Nanchang. Social bonding capital has a positive effect on the quality of life of PLWHA |
| 17 | Stakeholders' perspectives on integrating the management of depression into routine HIV care in Uganda: qualitative findings from a feasibility study | Conducted interviews with 11 PLWHA patients, doctors, supervisors, and mental health specialists and group-focused discussions by lay health workers. Interviews were conducted to evaluate HIV+D Interventions | Patients highlight the benefits of treating depression in the context of HIV care, including improved adherence to antiretroviral therapy, coping with sleeplessness, and suicidal ideation, and regaining a sense of selfefficacy. |
| 18 | Perceptions of Intersectional Stigma among Diverse Women Living with HIV in the United States | Conducted interviews with 76 women living with HIV from various socioeconomic backgrounds | Women living with HIV feel many forms of stigma apart from the stigma associated with HIV. The most common forms of stigma and discrimination are related to their gender, |
|  |  |  | race, and income level. |
| 19 | "My determination is to live": Narratives of African-American Women Who Have Lived with HIV for 10 or More Years | Conducting interviews with women living with HIV for 10 years | The need for specific programs for long-term survivors, including resilience training, educational programs on dating and disclosure, and opportunities to engage in meaningful work or volunteer initiatives |
| 20 | Learning to address multiple syndemics for people living with HIV through client perspectives on CBT | Conduct interviews with multiple comorbid PLWHA who complete cognitive-behavioral therapy (CBT) | Mental health issues are interrelated and provide an informative insight into how participants perceive the interconnections between their mental health diagnoses, and how they behave in their health. |

## 5. Discussion

The findings from this systematic review found that the perspective of PLWHA on stigma where stigma is still a major problem in improving their quality of life. Research by Jesus GJ, et al. (2017) the impact of living with a chronic disease that still carries a lot of stigma and prejudice is the biggest obstacle to PLWHA regarding their quality of life ^7^. It is difficult for PLWHA to improve their quality of life in community stigma that still exists today so that they do not receive social support in carrying out their lives. Through social support, especially family members, it is very important to help PLWHA accept their diagnosis and choose a healthy life and find ways to grow ^25^.

Many PLWHA expressed their perspective that it was difficult for them to be involved in a social environment because of the stigmatization of their HIV/AIDS status and made it difficult for them to seek social support from friends and family members. With the social conditions of PLWHA like that, they also lack emotional support so this can have an impact on their clinical condition. In this digital era, the development of information technology and social media makes PLWHA try to find alternatives in overcoming their social problems. PLWHA can seek support through online communities that they can access via the internet, where they can get information about their illness and find a community that suits them, so they can share experiences and their daily lives. In this way, PLWHA can get the social support that has an impact on their quality of life-related to their emotional needs. In the research of X. Han et al. (2018) with the development of social media in the PLWHA community, they can receive social support from online communities, where they can exchange information and share their experiences, as well as seek the needed emotional support ^27^.

In addition to the social support they seek through online communities, there is also a need for programs from the government that help PLWHA reduce the stigma they receive so that they can receive social support to improve their quality of life and their economic status. Several studies discuss the perspective of PLWHA about the need for programs to address social problems that occur. In the study of M. AbboahOffei et al. (2019) PLWHA understands patient-centered care (PCC) programs not only involve them in making decisions about their care but also as a way to deal with broader social problems such as living a normal life like other people, getting married, having children and being employed^16^

The limitations and weaknesses of this research lie in the process and systematic review, where the researcher realizes that in a study there must be many obstacles and obstacles. One of the factors that became an obstacle in this study was the limitation of language and time, where the researcher used only a literature review that used English where had an effect on the results obtained.

## 6. Conclusion

In this digital era, the perspective of PLWHA on stigma is still the main problem they face. However, with the development of technology and social media, they can get support from the online community they follow, so they can share experiences in improving their quality of life.

## Data Availability

All data produced in the present work are contained in the manuscript

https://journals.sagepub.com/

https://www.elsevier.com/solutions/sciencedirect

https://www.scopus.com

https://www.proquest.com

